# Transfer of Statistical Innovations of the 1990s-2000s in Oncology to the Biomedical Literature

**DOI:** 10.1101/19000638

**Authors:** Alexandre Vivot, Vincent Lévy, Sylvie Chevret

## Abstract

**Introduction:** Innovations in the fields of clinical studies require time to generate and disseminate new knowledge. We aimed to specifically explore lag times between the introduction and widespread use of innovative statistical methods in oncology using the competing risks and phase I model-based clinical trials settings as examples.

**Methods:** First, we defined a set of closed articles for each setting based on two princeps papers (Gray, Annals of Statistics 1998 for the competing risks setting and O’Quigley et al., Biometrics 1990 for the phase I setting). Secondly, we retrieved from the web of science all citations of the papers included in these sets. Each journal was classified as applied, semi-applied or methodological.

**Results:** A total of 6,727 citations for the competing risks setting and 2,639 citations for the phase I setting were found. Time to reach 25 citations was 6.2 years for the Gray’s paper and 4.5 years for the Fine and Gray paper, while it ranged from 3.4 years up to at least 20.1 years and not reached for 6 papers from the competing risks setting. The vast majority (91%) of the citing papers for the competing risks setting originated from applied journals. In contrast, less than half (44%) of the citing papers for the phase I setting were published in applied journals.

**Conclusion:** Statistical innovations in the competing risks setting have been widely diffused in the medical literature unlike the model-based designs for phase I trials, which are still seldom used 30 years after publication.

## Introduction

Translational medicine, aiming to expedite the discovery of new diagnostic tools and treatments by using a multi-disciplinary and collaborative approach, has grown interests in the last decade. Moreover, the commonly accepted delay of 17 years for research evidence to reach clinical practice recently appears to decrease [1] as exemplified by the mapping networks of publications and cross-references. [2] However, most translation gaps between knowledge and clinical application that have been investigated, notably in oncology, concerned translation of biological drivers into therapeutic benefits for patients. [3]

For the advancement of clinical research and eventually of patient care, the translation of innovative statistical methods into practice is also of crucial importance, though the average time elapsed for biostatical innovation to reach medical use is less known. In 1994, Altman showed that, while the key survival paper of Kaplan and Meier achieved only six citations in medical journals in the first 10 years after publication, evidence of decreasing lag times between the introduction and widespread use of innovative statistical methods was expected. [4] More than 2 decades later, we wondered whether the reported above decreasing dichotomy between basic or preclinical and clinical research [2] could be also true in the transfer of innovative statistical methods, confirming the Altman’s hypothesis. [4]

We first focused on competing risks methods, the key innovative survival methods of the last decades, [5] common in cancer. Indeed, competing risks exist whenever the probability of the main event (e.g., cancer) is prevented (e.g., by death) or altered (e.g., by transplantation) to occur. In this setting, standard survival methods that ignore the informative censoring of these competing events may result in biased effects of prognostic factors or treatments, [6] illustrating the need for bridging the gap of innovation into practice. Two innovative papers that first proposed a statistical test [7] and a regression model [8] for such data, were selected as the first innovative set.

With the aim of focusing on statistical innovations of potential large expected therapeutic benefits for patients, we secondly considered the setting of phase I clinical trials, which is also of great interest in the oncology area though with a limited diffusion as compared to phase III. [9] We chose as the most influential statistical pioneering approach, the continual reassessment method (CRM) that is, a model-based approach for dose-finding studies where only empiric 3+3 designs (formerly Fibonacci) were long available. We thus included in the second innovative set the original paper of the CRM [10] together with its close proposed modifications (Table 2).

**Table 1:**
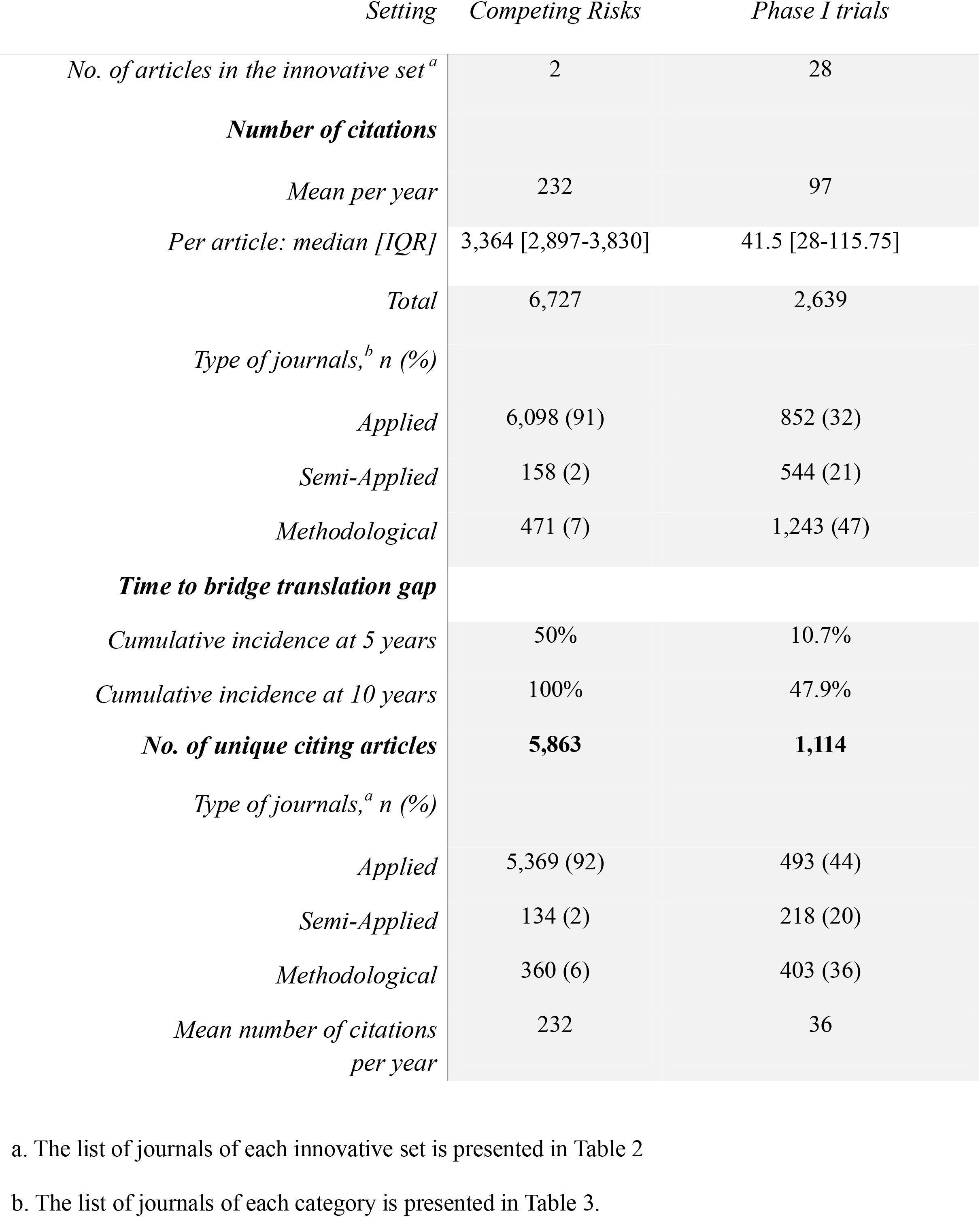
Number of citations retrieved from Web of Science on July 23, 2018

**Table 2.** List of selected articles for each innovative set.

| First Author | Journal | Pub. Year | No. of citations | Time to 25 citations (years) |
| --- | --- | --- | --- | --- |
| **Competing Risks Setting** |  |  |  |  |
| Fine | Journal of The American Statistical Association | 1999 | 4,296 | 4.5 |
| Gray | The Annals of Statistics | 1988 | 2,431 | 6.2 |
| **Phase I setting** |  |  |  |  |
| O'Quigley | Biometrics | 1990 | 676 | 5.7 |
| Goodman | Statistics In Medicine | 1995 | 258 | 5.0 |
| Babb J | Statistics In Medicine | 1998 | 235 | 5.3 |
| Cheung | Biometrics | 2000 | 181 | 3.4 |
| O'Quigley | Biometrics | 1996 | 170 | 7.1 |
| Korn | Statistics In Medicine | 1994 | 144 | 5.5 |
| Ratain | Journal of The National Cancer Institute | 1993 | 136 | 4.6 |
| Braun | Controlled Clinical Trials | 2002 | 109 | 5.6 |
| Moller | Statistics In Medicine | 1995 | 85 | 8.8 |
| Piantadosi | Cancer Chemotherapy And Pharmacology | 1998 | 84 | 10.4 |
| Chevret | Statistics In Medicine | 1993 | 68 | 10.3 |
| Whitehead | Journal of Biopharmaceutical Statistics | 1998 | 58 | 9.8 |
| Mick | Journal of The National Cancer Institute | 1993 | 46 | 8.4 |
| Tighiouart | Statistics In Medicine | 2005 | 42 | 9.5 |
| O'Quigley | Biometrics | 1992 | 41 | 9.5 |
| Reiner | Computational Statistics & Data Analysis | 1999 | 38 | 13.2 |
| Ishizuka | Statistics In Medicine | 2001 | 37 | 10.9 |
| O'Quigley | Biostatistics | 2002 | 36 | 11.8 |
| O'Quigley | Biometrics | 2003 | 35 | 11.8 |
| O'Quigley | Journal of Biopharmaceutical Statistics | 1999 | 32 | 16.6 |
| Babb | Statistics In Medicine | 2001 | 29 | 15.6 |
| Zacks | Statistics & Probability Letters | 1998 | 25 | 20.1 |
| Rogatko | Clinical Cancer Research | 2005 | 19 | Not reached |
| O'Quigley | Journal of Statistical Planning And Inference | 2006 | 17 | Not reached |
| Doussau | Statistics In Medicine | 2013 | 14 | Not reached |
| Legedza | Controlled Clinical Trials | 2000 | 10 | Not reached |
| Shu | Statistics In Medicine | 2008 | 7 | Not reached |
| Mahmood | Journal of Clinical Pharmacology | 2001 | 7 | Not reached |

The primary objective of this paper was to assess whether Altman’s prediction was fulfilled in the field of oncology using the competing risks and the phase I clinical trials as examples. Secondary objectives were to study the relative importance of the applied and statistical literature in papers citing statistical innovations and to analyze which medical areas the methods are mainly used.

## Methods

### Selection of articles

Based on our knowledge, we *a priori* included the paper by Gray[7] and that by Fine and Gray [8] in the innovative set for the competing risks setting. For the phase I setting, beside the paper by O’Quigley, [10] because numerous modifications of the initial CRM very similar to the initial paper were published, we jointly considered the set of those articles. Thus, we retrieved all papers citing O’Quigley’s paper [10] in the Web of Science on May 9, 2018. We then constructed a co-citations network, where co-citations were defined as links between two papers, both of which cited by the same paper. The relatedness of papers was based on the number of times they have been cited together. This process was used to create the innovative set for the phase I trials setting.

### Citation data

For both settings, we retrieved all citations of the innovative set from the Web of Science on July, 23 2018. Citation data were imported into R software (https://www.R-project.org/) using the Bibliometrix package. [12] The information retrieved included the title, abstract, date of publication, journal name and category according to the classification by Incites Journal Citation (https://jcr.incites.thomsonreuters.com). We excluded animal studies by searching animal names (cat, dog, mice, mouse, rat, monkey, primate, macaque) in the title. Each citation article was also segregated to one of the three following categories according to the journal of publication: 1) applied journals, 2) semi-applied journals, and 3) methodological journals (Table 3). For citations of the phase I setting, one investigator (AV) manually reviewed the papers with “phase I” in title to check if they were actual dose-finding phase I clinical trials.

**Table 3.**
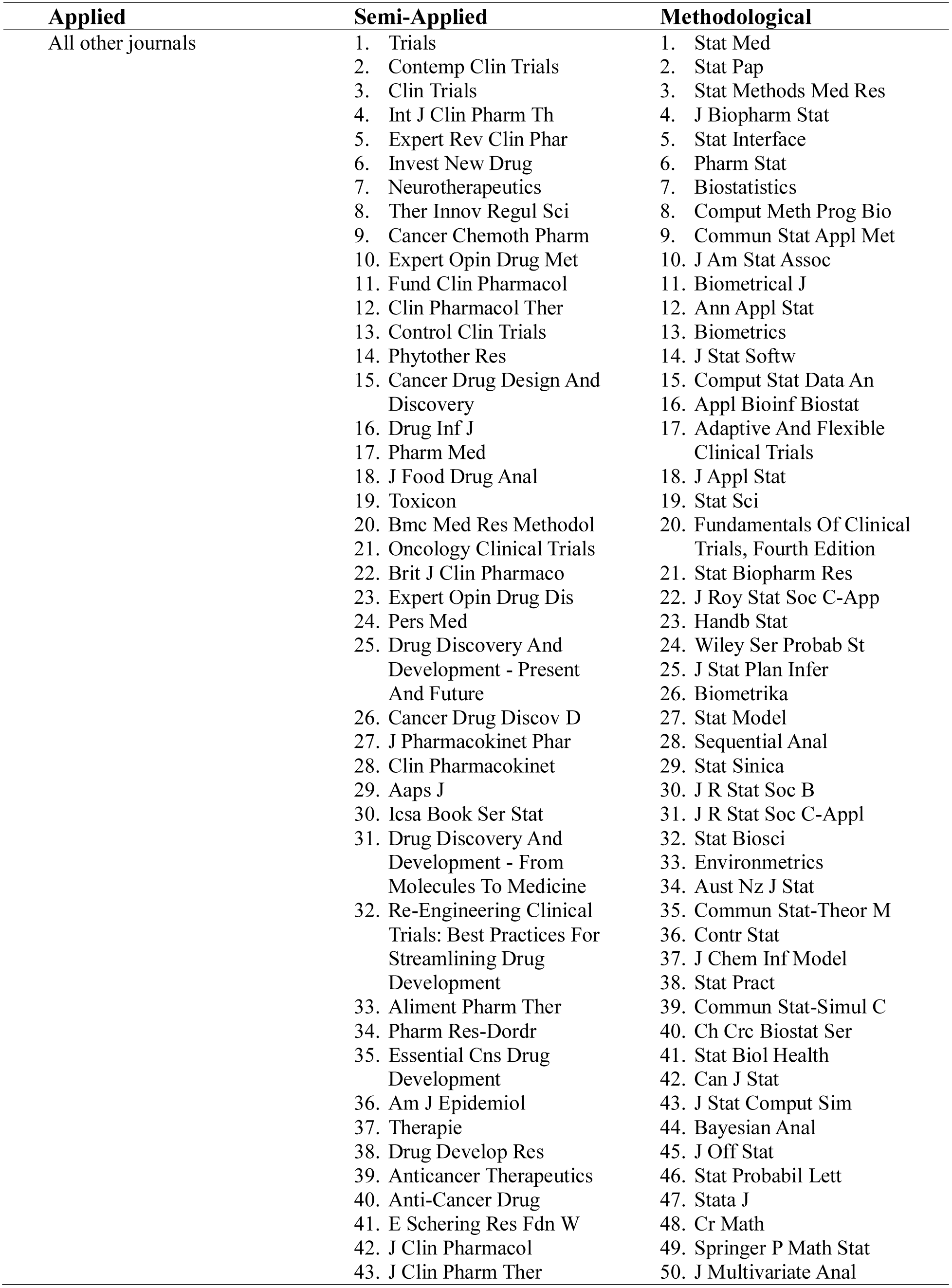
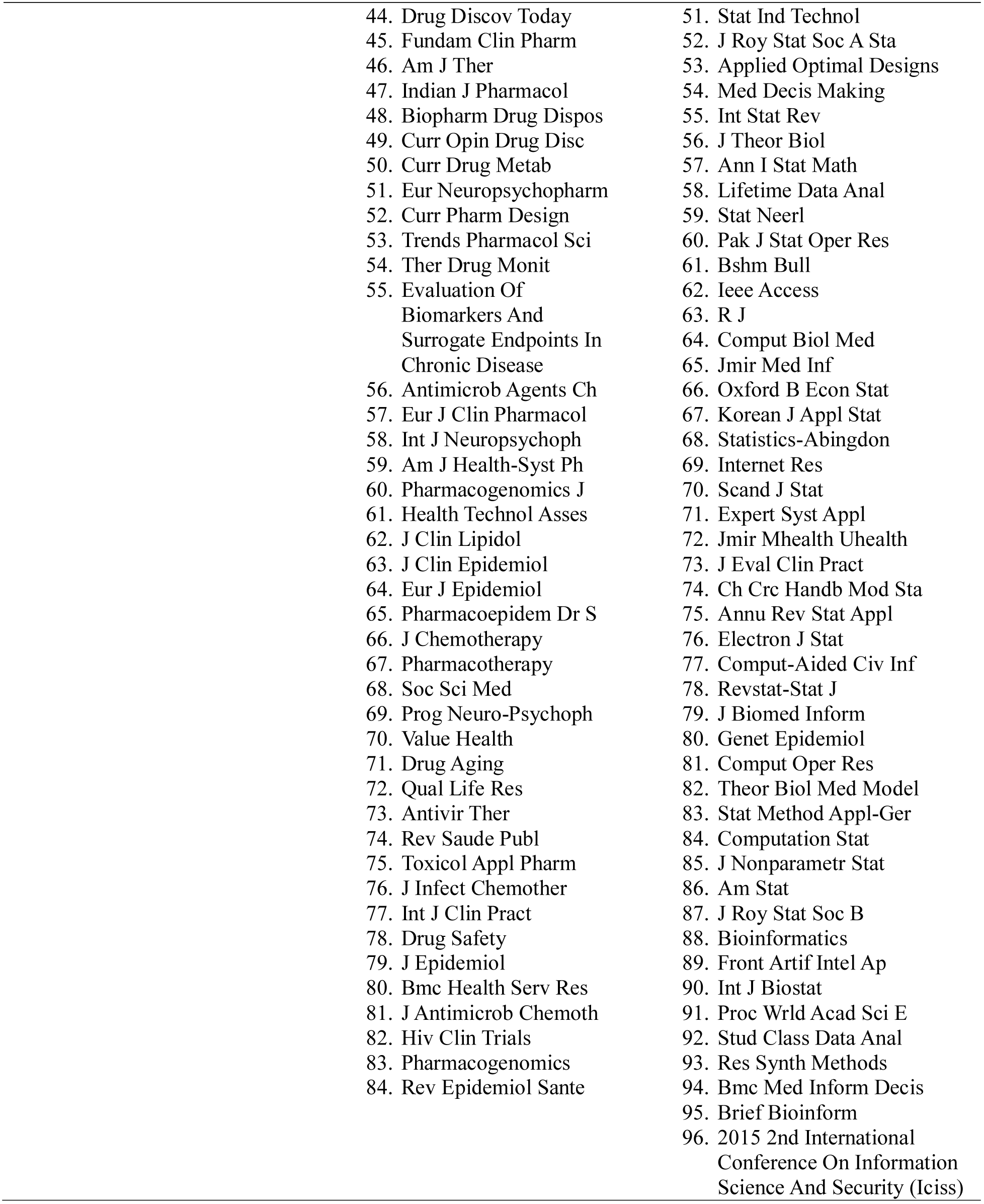
List of selected journals for each class.

| **Applied** | **Semi-Applied** | **Methodological** |
| --- | --- | --- |
| All other journals | 1. Trials<br>2. Contemp Clin Trials<br>3. Clin Trials<br>4. Int J Clin Pharm Th<br>5. Expert Rev Clin Phar<br>6. Invest New Drug<br>7. Neurotherapeutics<br>8. Ther Innov Regul Sci<br>9. Cancer Chemoth Pharm<br>10. Expert Opin Drug Met<br>11. Fund Clin Pharmacol<br>12. Clin Pharmacol Ther<br>13. Control Clin Trials<br>14. Phytother Res<br>15. Cancer Drug Design And Discovery<br>16. Drug Inf J<br>17. Pharm Med<br>18. J Food Drug Anal<br>19. Toxicon<br>20. BMC Med Res Methodol<br>21. Oncology Clinical Trials<br>22. Brit J Clin Pharmacol<br>23. Expert Opin Drug Dis<br>24. Pers Med<br>25. Drug Discovery And Development - Present And Future<br>26. Cancer Drug Discov D<br>27. J Pharmacokinet Phar<br>28. Clin Pharmacokinet<br>29. Aaps J<br>30. ICSA Book Ser Stat<br>31. Drug Discovery And Development - From Molecules To Medicine<br>32. Re-Engineering Clinical Trials: Best Practices For Streamlining Drug Development<br>33. Aliment Pharm Ther<br>34. Pharm Res-Dordr<br>35. Essential CNS Drug Development<br>36. Am J Epidemiol<br>37. Therapie<br>38. Drug Develop Res<br>39. Anticancer Therapeutics<br>40. Anti-Cancer Drug<br>41. E Schering Res Fdn W<br>42. J Clin Pharmacol<br>43. J Clin Pharm Ther | 1. Stat Med<br>2. Stat Pap<br>3. Stat Methods Med Res<br>4. J Biopharm Stat<br>5. Stat Interface<br>6. Pharm Stat<br>7. Biostatistics<br>8. Comput Meth Prog Bio<br>9. Commun Stat Appl Met<br>10. J Am Stat Assoc<br>11. Biometrical J<br>12. Ann Appl Stat<br>13. Biometrics<br>14. J Stat Softw<br>15. Comput Stat Data An<br>16. Appl Bioinf Biostat<br>17. Adaptive And Flexible Clinical Trials<br>18. J Appl Stat<br>19. Stat Sci<br>20. Fundamentals Of Clinical Trials, Fourth Edition<br>21. Stat Biopharm Res<br>22. J Roy Stat Soc C-App<br>23. Handb Stat<br>24. Wiley Ser Probab St<br>25. J Stat Plan Infer<br>26. Biometrika<br>27. Stat Model<br>28. Sequential Anal<br>29. Stat Sinica<br>30. J R Stat Soc B<br>31. J R Stat Soc C-App<br>32. Stat Biosci<br>33. Environmetrics<br>34. Aust Nz J Stat<br>35. Commun Stat-Theor M<br>36. Contr Stat<br>37. J Chem Inf Model<br>38. Stat Pract<br>39. Commun Stat-Simul C<br>40. Ch Crc Biostat Ser<br>41. Stat Biol Health<br>42. Can J Stat<br>43. J Stat Comput Sim<br>44. Bayesian Anal<br>45. J Off Stat<br>46. Stat Probabil Lett<br>47. Stata J<br>48. Cr Math<br>49. Springer P Math Stat<br>50. J Multivariate Anal |

|  |  |
| --- | --- |
| 44. Drug Discov Today | 51. Stat Ind Technol |
| 45. Fundam Clin Pharm | 52. J Roy Stat Soc A Sta |
| 46. Am J Ther | 53. Applied Optimal Designs |
| 47. Indian J Pharmacol | 54. Med Decis Making |
| 48. Biopharm Drug Dispos | 55. Int Stat Rev |
| 49. Curr Opin Drug Disc | 56. J Theor Biol |
| 50. Curr Drug Metab | 57. Ann I Stat Math |
| 51. Eur Neuropsychopharm | 58. Lifetime Data Anal |
| 52. Curr Pharm Design | 59. Stat Neerl |
| 53. Trends Pharmacol Sci | 60. Pak J Stat Oper Res |
| 54. Ther Drug Monit | 61. Bshm Bull |
| 55. Evaluation Of<br>Biomarkers And<br>Surrogate Endpoints In<br>Chronic Disease | 62. Ieee Access |
| 56. Antimicrob Agents Ch | 63. R J |
| 57. Eur J Clin Pharmacol | 64. Comput Biol Med |
| 58. Int J Neuropsychoph | 65. Jmir Med Inf |
| 59. Am J Health-Syst Ph | 66. Oxford B Econ Stat |
| 60. Pharmacogenomics J | 67. Korean J Appl Stat |
| 61. Health Technol Asses | 68. Statistics-Abingdon |
| 62. J Clin Lipidol | 69. Internet Res |
| 63. J Clin Epidemiol | 70. Scand J Stat |
| 64. Eur J Epidemiol | 71. Expert Syst Appl |
| 65. Pharmacoepidem Dr S | 72. Jmir Mhealth Uhealth |
| 66. J Chemotherapy | 73. J Eval Clin Pract |
| 67. Pharmacotherapy | 74. Ch Crc Handb Mod Sta |
| 68. Soc Sci Med | 75. Annu Rev Stat Appl |
| 69. Prog Neuro-Psychoph | 76. Electron J Stat |
| 70. Value Health | 77. Comput-Aided Civ Inf |
| 71. Drug Aging | 78. Revstat-Stat J |
| 72. Qual Life Res | 79. J Biomed Inform |
| 73. Antivir Ther | 80. Genet Epidemiol |
| 74. Rev Saude Publ | 81. Comput Oper Res |
| 75. Toxicol Appl Pharm | 82. Theor Biol Med Model |
| 76. J Infect Chemother | 83. Stat Method Appl-Ger |
| 77. Int J Clin Pract | 84. Computation Stat |
| 78. Drug Safety | 85. J Nonparametr Stat |
| 79. J Epidemiol | 86. Am Stat |
| 80. BMC Health Serv Res | 87. J Roy Stat Soc B |
| 81. J Antimicrob Chemoth | 88. Bioinformatics |
| 82. Hiv Clin Trials | 89. Front Artif Intel Ap |
| 83. Pharmacogenomics | 90. Int J Biostat |
| 84. Rev Epidemiol Sante | 91. Proc Wrld Acad Sci E |
|  | 92. Stud Class Data Anal |
|  | 93. Res Synth Methods |
|  | 94. BMC Med Inform Decis |
|  | 95. Brief Bioinform |
|  | 96. 2015 2nd International<br>Conference On Information<br>Science And Security (Iciss) |

### Statistical analysis

Analysis was performed for each setting, separately. Summary statistics were reported, either mean (standard deviation) or median [interquartile range]. We plotted the cumulative number and proportions of “applied”, “semi-applied”, and “methodological” fractions of citations ^5^ over time as a percentage of the citations classified in the three categories of journals defined above. We defined translational gap as the time between the publication of the biostatistical paper and the time it reached 25 citations in applied literature, as defined by Altman; [4] estimated cumulative incidence of translational gap was estimated by the Kaplan Meier approach, due to the administrative censoring of the data on 2018.

To obtain further insight into the medical literature citing these innovative statistical tools, we constructed a citation network using the journal as the statistical unit (rather than the paper). Use of this network, together with clustering techniques, allowed us to analyze which medical areas (using the journal as a proxy) cited the most statistical innovations. VosViewer was used to create maps and clustering analysis. [14]

### Ethical Statement

All methods were carried out in accordance with relevant guidelines and regulations. No informed consent or ethics approval was necessary because this study is based on publicly available data and involved no individual patient data collection or analysis.

## Results

The innovative sets of papers used for both settings are reported in Table 2. They included 2 articles for the competing risks set, and 28 for the phase I set.

### Competing risks set

A total of 6,727 citations (5,863 unique articles), including 2,431 for Gray’s article and 4,296 for the Fine and Gray paper, were found (Table 1). Time to translational gap was 6.2 years for the Gray’s paper and 4.5 years for the Fine and Gray paper. The vast majority (91%) of the citations originated from applied journals, with a sharp and continuous increase in citations over time (Figure 1); in contrast, the citations from methodological and semi-applied journals represented only small percentages (6% and 2%, respectively) of the citing papers over the entire period. This was confirmed by the representation of the network of these journals (Figure 2).

**Figure 1.**
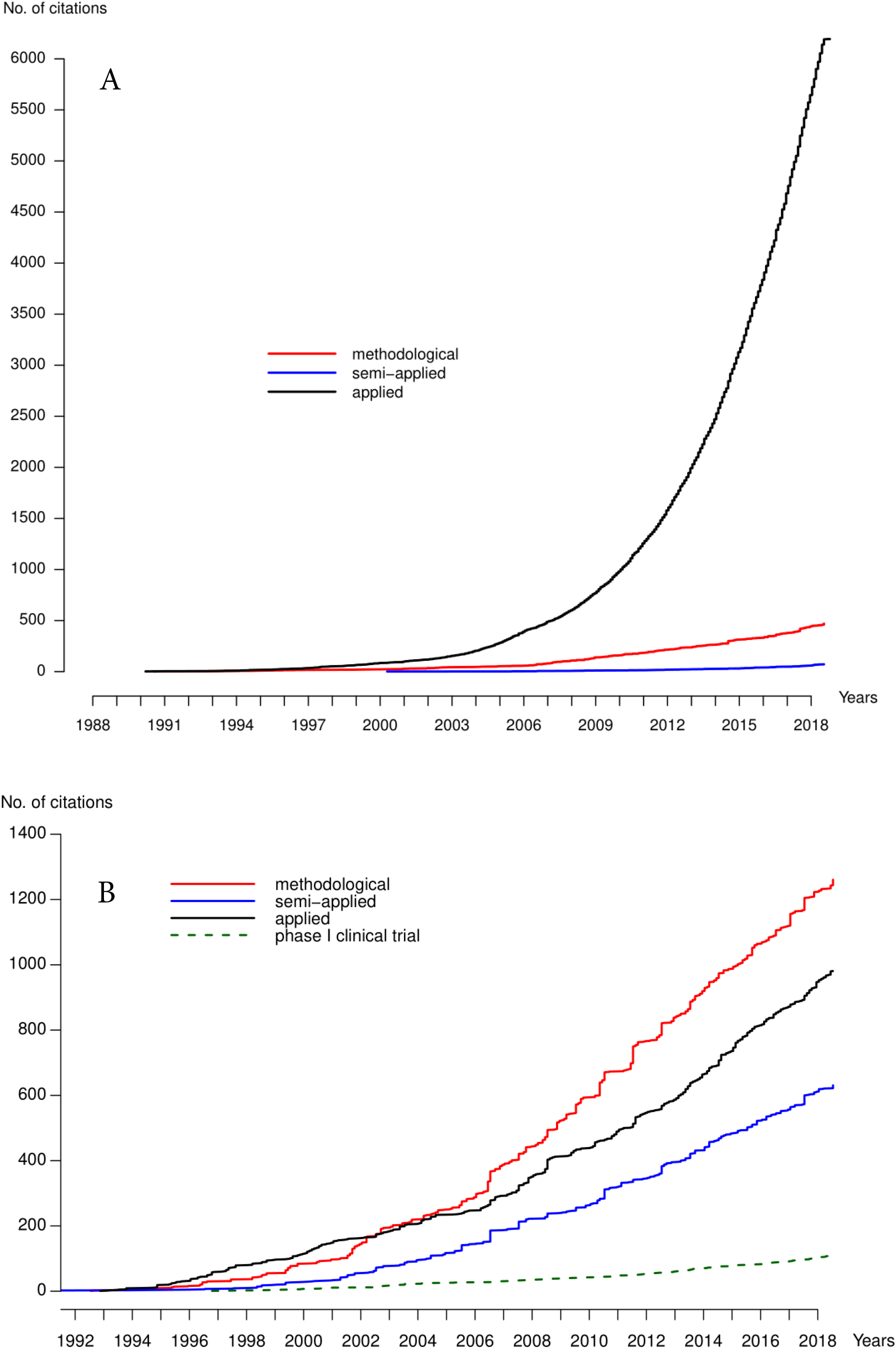
Cumulative number of citations for the competing risks setting (Panel A) and the phase I trials setting (Panel B) retrieved from the Web of Science and classified in each category of journals.

**Figure 2.**
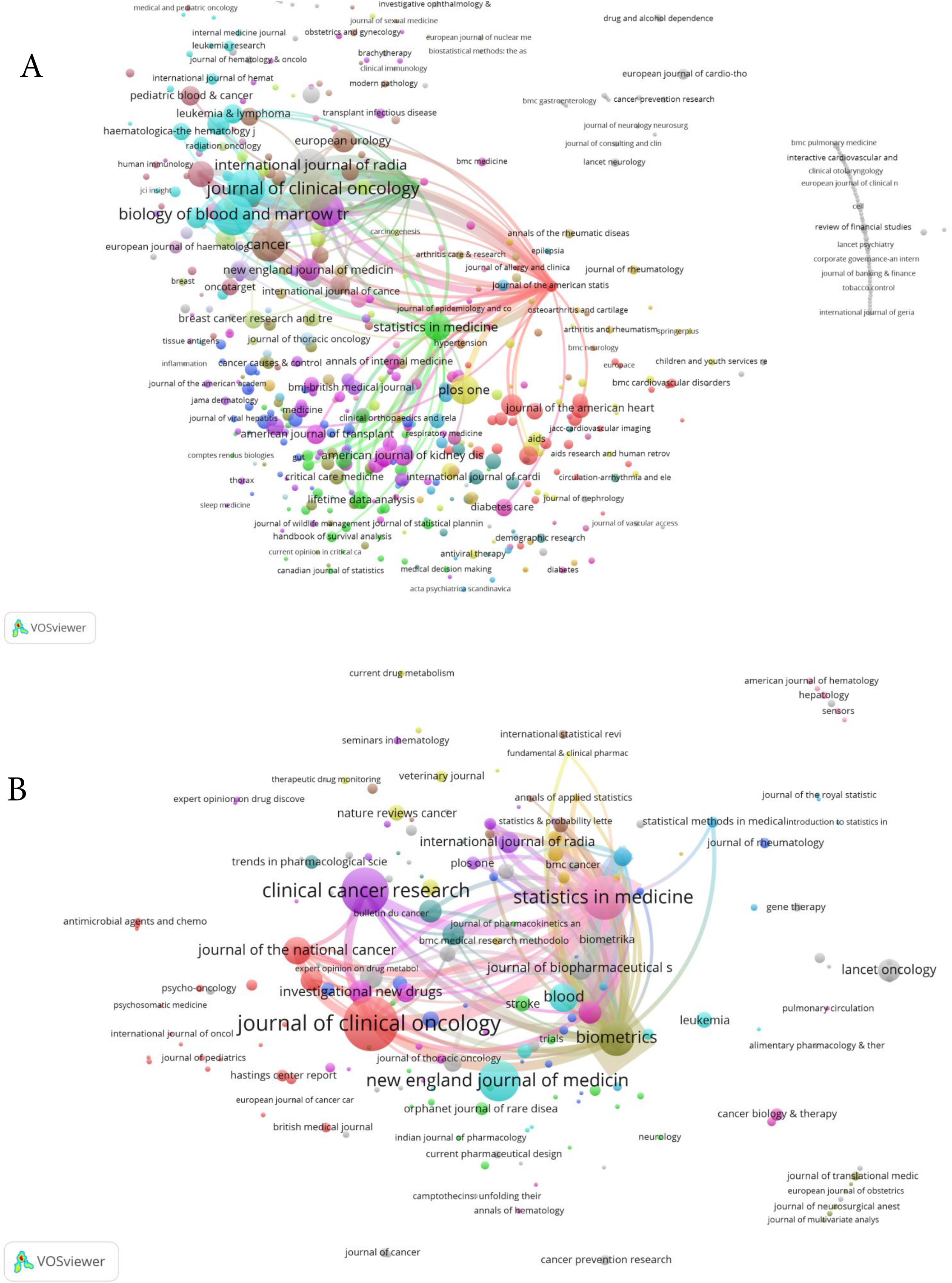
Citation network of journals for the two innovative sets (Competing risks- Fig 2A, Phase I trials- Fig 2B). For clarity, only journals with at least 5 citations are represented.

Beside a methodological cluster, two major clusters confirmed the two main areas of application of competing risks, namely oncology (e.g., Journal of Clinical Oncology, European Journal of Cancer and Hematology) and hematology (e.g., Blood, Haematologica, and Bone Marrow Transplantation). Other clusters represent other areas of application, such as cardiovascular diseases.

### Phase I clinical trials set

After excluding books and animal studies, a total of 2,639 citations (1,114 unique papers after removing duplicates) for the phase I innovative set were found. The three most cited papers were published by O’Quigley in 1999 (676 citations), Goodman in 1995 (258 citations) and Babb in 1998 (235 citations). Contrarily to the competing risks setting, the translational gap differed across papers, from 3.4 years up to at least 20.1 years since not reached for 6 papers. Overall, it was reached by 10.7% of the set articles at year-5 and by 47.9% at year-10. Less than one half (44%) of citing articles were published in applied journals, more than one third (36%) in statistics journals, and a fifth (20%) in semi-applied journals, with roughly similar rate of citations over time (Figure 1). When we restricted the citations to those articles with “phase I” in the title, only 415 (37%) articles were selected; of these, only 110 were found to be phase I clinical trials after manual reviewing the titles.

Network of the journals of papers citing one of the phase I set is displayed in Figure 2. The right part of the graph represents the methodological cluster containing, for example, Statistics in Medicine and Biometrics. The left part of the graph represents the large group concerning applications or reviews of the CRM in the most important clinical cancer journals (Journal of Clinical Oncology, Clinical Cancer Research, Annals of Oncology, British Journal of Cancer, and Journal of National Cancer Institute); in contrast, non-cancer journals had published only a few studies.

## Discussion

This study aimed to check whether the time lags in the statistical translation process was actually shortened, focusing on two areas with a large potential for clinical research improvements and widely encountered in the oncology literature, namely, the survival methods for competing risks data and the new designs for phase I clinical trials. The answer was bifid, with a short translational gap, defined by the time to reach 25 citations, [4] of about 5 years for the former, but delayed above 10 years for one half of the later. This could be expected given the two statistical innovative sets are used at different stages of clinical studies, with different levels of complexity. Indeed, the first set of competing risks articles deals with method of data analysis, that can easily be performed using modern statistical software without the need for major expertise - although whether such studies are used, performed and interpreted correctly can be another matter. [15] Therefore, such a lag time of about 5 years for using an innovative data analytic method is in agreement with the time to publication after completion of data collection and analysis recently estimated at about 3 years in six journals with high impact factors. [16] In contrast, the second set of innovative methods concern a change in trial design and logistics; thus, beside the time of analysis and publication, the transfer of innovative clinical trial design into the medical literature is obviously impacted by the additional constraints of patient enrollment time, and the follow-up period for the end point. It moreover requires statisticians to engage since the planning phase, [17] up to the analysis of the data, contrary to traditional methods such as the ‘3+3’ design, which can be used without involving statisticians and computer programs and remains the most common choice among clinicians for phase I dose-escalation oncology trials. [18]

Moreover, difference in complexity of both settings was illustrated in terms of citation patterns. Although the competing risks methodology was widely diffused over the medical (in particular, oncology) community, methodology relating to innovative designs for cancer Phase I trials failed to translate easily into practice, consistent with the results of a previous study that showed a very slow transfer of phase I design improvements into clinical practice. [19]

Nevertheless, the two settings share large implications for clinical research, and researchers trying to apply the statistical innovative methods should not be delayed in using the new knowledge. This is notably true in the setting of phase I trials in oncology, where improved selection of patients due to improvements in translational medicine should translate into faster and more precise dose determination. [20] Diffusing more widely into the applied literature and the medical community could be achieved in several ways. First, communication between biostatisticians and clinical colleagues should be improved, although many reviews have been published on these methods in the medical literature, as illustrated by the non-negligible fraction of semi-applied and applied papers that are different from the original clinical trials used in our study. This could be driven through key gateway journals, as suggested by the clusters of citation journals where Journal of Clinical Oncology and Statistics in Medicine appear major players (Figure 2). Concerning competing risks analysis, for which adequate modern methods have been integrated into all modern statistical software, a key step may be to educate clinicians to recognize the settings in which competing risks are of concern and to persuade them of the importance of using those methods. For the design of phase I studies, improvements in providing concrete guidance for designing such trials and facilitating their implementation in practice is still mandatory for bridging the gap between statistical innovation and practical implementation. Model-based designs are cited in the US FDA guidance for industries that are related to adaptive clinical trials as “less understood models,” and the FDA highlights some of their disadvantages. [21]

Our study has some limitations. First, we used citations as a measure of the transfer of knowledge between researchers and physicians. Such citation counts, although not a direct measure of the intrinsic significance of a research idea, provide a measure of statistical technology transfer and of its impact. [4] However, in addition to papers that focus on ranking journals and impact factors [22] many others have considered the citations only qualitatively. [23] Second, we segregated articles into three categories of applied, semi-applied and methodological journals, as previously reported. [5] However, this distinction is somewhat simplistic because the applied fractions of articles published in statistical journals may increase over time; this is reportedly highest for statistical medicine. [5] Third, we focused on two main and distinct topics of biostatistics, and the results might differ for other statistical innovations, such as dynamic prediction modelling, the joint modelling of longitudinal data, and multiple imputation techniques for handling missing data.

### Conclusion

In summary, statistical innovations for the competing risks setting have been widely diffused in the medical literature, especially in oncology and hematology, fulfilling Altman’s prediction about decreasing lag times, unlike the model-based designs for phase I trials, which are still seldom used 30 years after their first publication. However, for both statistical methods, a translational gap remains that needs to be filled before the oncology community can benefit fully from these modern methods.

## Data Availability

Data used in this study are publicly available data. The datasets generated and analyzed during the current study are available from the corresponding author on reasonable request.

## Abbreviations

CRM: Continual Reassessment Method

## Authors’ contributions

SC had the original idea. SC and AV designed the study. AV conducted the literature search, extracted and analyzed data. AV, VL, and SC participated in the interpretation of the data. AV drafted the manuscript. AV, VL, and SC authors reviewed the manuscript and approved the final version.

## Acknowledgements

We wish to thank Dr Lucie Biard for helpful comments on the manuscript. We acknowledge the paid contribution of American Journal Experts (Research Square, Durham, NC, USA) for copyediting.

